# Thyroid function and risk of anemia: a multivariable-adjusted and Mendelian Randomization analysis in the UK Biobank

**DOI:** 10.1101/2021.06.24.21259436

**Authors:** Nicolien A. van Vliet, Annelies E.P. Kamphuis, Wendy P.J. den Elzen, Gerard J. Blauw, Jacobijn Gussekloo, Raymond Noordam, Diana van Heemst

**Affiliations:** Department of Internal Medicine, Section of Gerontology and Geriatrics, Leiden University Medical Center, Leiden, the Netherlands; Atalmedial Diagnostics Centre, Amsterdam the Netherlands; Department of Clinical Chemistry, Amsterdam UMC, Amsterdam, the Netherlands; Department of Public Health and Primary Care, Leiden University Medical Center, Leiden, the Netherlands

**Keywords:** Thyroid status, Anemia, Mendelian Randomization, UK Biobank

## Abstract

**Context:** Thyroid dysfunction is associated with higher anemia prevalence, though causality remains unclear.

**Objective:** To investigate a potential causal relationship between thyroid function and anemia.

**Design:** Cross-sectional and Mendelian Randomization study

**Setting:** Europeans from UK Biobank

**Participants:** 445,482 participants, mean age 56.77 years (SD 8.0) and 54.2% were women. Self-reported clinical diagnosis of hypothyroidism was stated by 21,860 (4.9%); self-reported clinical diagnosis of hyperthyroidism by 3,431 (0.8%).

**Main Outcome Measure:** Anemia, defined as hemoglobin level of <13 g/dL in men and <12 g/dL in women, was present in 18,717 (4.2%) participants.

**Results:** In cross-sectional logistic regression analyses, self-reported clinical diagnoses of hypo- and hyperthyroidism were associated with higher odds of anemia (OR 1.12, 95%CI 1.05-1.19 and OR 1.09, 95%CI 0.91-1.30), though with wide confidence intervals for hyperthyroidism. Although we considered a possible non-linear relationship, we did not observe an association of higher or lower genetically-influenced thyrotropin (TSH) with anemia (versus middle tertile: OR lowest tertile 0.98, 95% CI 0.95-1.02; highest tertile 1.02, 95% CI 0.98-1.06), nor of genetically-influenced free thyroxine (fT4) with anemia. Individuals with genetic variants in the *DIO3OS* gene implicated in intracellular regulation of thyroid hormones had a higher anemia risk (OR 1.05, 95% CI 1.02-1.10); no association was observed with variants in *DIO1* or *DIO2* genes.

**Conclusions:** While self-reported clinical diagnosis of hypothyroidism was associated with a higher prevalence of anemia, we did not found evidence supporting this association was causal. However, intracellular regulation of thyroid hormones might play a role in developing anemia.

## Introduction

Anemia is a very common condition, affecting 27% of the world population (1). The definition of anemia is a deficiency of healthy erythrocytes, associated with reduced circulating levels of hemoglobin (Hb); the World Health Organization set a universal threshold for anemia at a Hb level of <13g/dL in men and <12g/dL in women (2). Common causes of anemia include deficiency of iron, vitamin B12 and folate, chronic kidney disease, and inflammatory diseases (3,4). One such other possible cause for the development of anemia is an abnormal thyroid status (5).

Regulation of thyroid hormone availability is complex and occurs both centrally (HPT-axis) and locally (by differential expression of thyroid hormone transporters, receptors and deiodinases) (6). Under physiological conditions, both circulating thyroid stimulating hormone (TSH) and free thyroxine (fT4) concentrations are regulated through negative-feedback by the Hypothalamus-Pituitary-Thyroid (HPT) axis. In target tissues, deiodinases can activate or deactivate thyroid hormones: deiodinase type 1 and 2 (D1 and D2) can convert the prohormone T4 into the active hormone triiodothyronine (T3), while deiodinase type 3 (D3) can convert T4 into inactive reverse T3 (7). The central regulation may fail in presence of thyroid gland pathology, causing either hypothyroidism (biochemically characterized by low fT4 and elevated TSH) (8) or hyperthyroidism (characterized by elevated fT4 and low TSH) (9).

Multiple studies have suggested an association between thyroid dysfunction and anemia, including a possible U-shaped relationship between thyroid hormone levels and anemia (10), but causality of these associations remain to be proven (10-12). To investigate whether the relationship between thyroid dysfunction and anemia is causal, we used a multi-approach strategy and performed cross-sectional multivariable-adjusted logistic regression, Mendelian Randomization (MR) and weighted candidate gene analyses. Multivariable-adjusted regression analyses were conducted to replicate previous studies regarding the association of hypo-and hyperthyroidism and anemia. In MR, genetic variants associated with the exposure are used as instrumental variables free from most confounding and reverse causation (13). In the present MR study, we used genetic instruments for TSH and fT4 that were obtained in populations with participants within the euthyroid range as exposure, and anemia the outcome. For the weighted candidate gene analysis, we used genetic variants mapped to genes encoding deiodinases which were identified in recent genome-wide association studies (GWAS) (14) as associated with circulating fT4 levels. We therefore hypothesized that these variants may have functional implications in local thyroid hormone homeostasis.

## Methods

### Study population

The UK Biobank (UKB) cohort is a prospective general population cohort. Baseline assessments took place between 2006 and 2010 in 22 different assessment centers across the United Kingdom (15). A total of 502,628 participants aged between 40 and 70 years were recruited from the general population. Invitation letters were sent to eligible adults registered to the National Health Services (NHS) and living within a 25 miles distance from one of the study assessment centers. The response rate was 5.5% (16). At the study assessment center, participants completed questionnaires through touchscreen that included topics as sociodemographic characteristics, physical and mental health, lifestyle and habitual food intake.

For the present study, we selected all individuals with a self-reported European ancestry, available genomics data and data on Hb levels. As a result, we included a total sample of 445,482 individuals. The UK Biobank has approval from the NHS North West Multi-Centre Research Ethics Committee (ref 11/NW/0382). All participants from the UK Biobank cohort provided written informed consent, and the study was approved by the medical ethics committee. The current project was completed under project number 32743.

### Exposure assessment

#### Self-reported clinical diagnosis of hypo-or hyperthyroidism

At the assessment centre, UKB participants were asked in a verbal interview by a trained research nurse whether they suffered from either a clinical diagnosis of hypo-or hyperthyroidism, and whether they used any medication for thyroid dysfunction (15). Within this study, liothyronine (T3 suppletion), levothyroxine (T4 suppletion), anti-thyroid medication or a combination therapy were considered as thyroid medication (ATC code H03).

#### Selection of the genetic instruments for TSH and fT4 for the Mendelian Randomization

We selected all independently associated single nucleotide polymorphisms (SNPs) (*p* value <5×10^−8^ (13)) for circulating TSH and fT4 levels within the reference range as instrumental variables from a previously performed genome-wide association meta-analysis (14). Study-specific protocols of the studies contributing to the meta-analysis are described previously (14). This genome-wide association meta-analysis, being the largest conducted in European-ancestry participants on TSH and fT4 concentrations in the euthyroid range, was performed in 71,167 European-ancestry participants from 22 different cohorts (14). From this effort, we derived a total of 62 SNPs associated with TSH (explaining 9.4% of the total variance), and 31 SNPs associated with fT4 (explaining 4.8% of the total variance). Based on the SNP effect sizes, we calculated weighted genetic risk scores for each individual included in the study.

#### Genetic risk scores based on deiodinase activity

To explore the role of deiodinase activity in anemia implicated in intracellular regulation of thyroid hormones, we calculated separate genetic risk scores based on fT4-associated SNPs mapped to either *DIO1, DIO2*, and *DIO3OS*. These scores were weighted to the association of each SNP with circulating fT4, as a surrogate for the magnitude of the intracellular effects.

### Anemia

For measuring blood Hb levels, blood was collected into a 4-mL EDTA vacutainer and held in temperature-controlled shipping boxes (4°C). Complete blood cell counts were conducted using a Coulter Counter. The universal definition of anemia in adults is an Hb level of <13 g/dL in men and <12 g/dL in women, as proposed by the WHO (2).

### Covariates

Low-grade systemic inflammation was considered a possible important confounder for the analyses with self-reported clinical diagnosis of hypo-and hyperthyroidism, since inflammation affects both thyroid status (17) and risk of anemia (18) via distinct biological pathways. These analyses were therefore adjusted for factors related to low-grade systemic inflammation, notably levels of high sensitive C-reactive protein (hs-CRP) and the lifestyle factors smoking and alcohol intake. Smoking status was assessed via a touchscreen questionnaire inquiring whether the participant was currently smoking, past smoker or never smoker; for the present study we dichotomized smoking status into current smoker or non-smoker. Alcohol intake was also assessed by touchscreen questionnaire. All participants were asked how frequently they drank alcoholic beverages. As there is no guideline on frequency of drinking alcoholic beverages, we aimed to divide into two groups of similar size which could be interpreted as drinking more or less frequently than average. For the present population the cut-off point was up to two times per week or more than twice a week.

### Statistical analyses

Participant characteristics were presented in the whole study population as well as in men and women separately as mean (standard deviation; for normally distributed continuous data), as median (interquartile range; for non-normal distributed data) or as proportion (for categorical variables).

We first performed multivariable-adjusted logistic regression analyses to investigate the associations between self-reported hypo-and hyperthyroidism and anemia. Two models were constructed; a minimally-adjusted model comprising age and sex, and a fully-adjusted model additionally comprising natural log-transformed hs-CRP, smoking status and alcohol intake. For sensitivity purposes, the dichotomous outcome of anemia was supplemented with analyses of Hb as a continuous outcome. Similar models were constructed as above, but applied to multivariable-adjusted linear regression to assess the difference in Hb in g/dL.

Subsequently, we assessed the association of genetically-influenced TSH and fT4 concentration, as instrumental variables, with anemia in the total study population. Since the previously found association, based on a large multi-cohort analysis, between thyroid function and anemia by Wopereis *et al*. was U-shaped (10), the genetic risk scores were divided into equally-sized tertiles. With the middle group as reference, a multivariable-adjusted logistic regression model was constructed adjusted for age, sex and the first 10 principal components to correct for possible population stratification. Similar models were built for the genetic risk scores for deiodinase activity, again with the middle tertile as a reference group. To assess consistency, similar models were constructed to investigate the difference in Hb in g/dL between the highest and lowest tertile of genetic risk scores compared to the middle tertile using multivariable-adjusted linear regression.

To assess sex-specific associations, the main analyses were also performed stratified by sex and interaction analyses (on a multiplicative scale) were performed when sex differences were apparent by including an interaction term between sex and the exposure in the logistic regression model. In addition, summary-level data-based methods for MR were employed to assess whether unbalanced directional pleiotropy may have biased our analyses on genetically-determined TSH and fT4 with anemia. The inverse variance-weighted (IVW) analysis provides a weighted mean estimate of the association of the individual genetic variants which assumes that none of the instruments were invalid(19). In addition, the weighted median estimator (WME), MR Egger regression and MR pleiotropy residual sum and outlier (MR-PRESSO) analyses were employed as they do take into account possible bias caused by directional pleiotropy based on different assumptions on the number of invalid instrumental variables (19,20).

The results for the multivariable logistic regression of self-reported hypo-and hyperthyroidism are presented as the estimated Odds Ratio (OR) of having anemia for those who reported hypo-or hyperthyroidism compared to those without thyroid disease with the accompanying 95% confidence interval (95% CI). For the multivariable logistic regression of tertiles of genetic risk score, the OR and 95% CI of having anemia are given for the 33% with the highest and the lowest genetically determined TSH and fT4 compared to the middle third. For the multivariable linear regression analyses, we present the estimated difference in Hb concentration (expressed in g/dL) between the groups with accompanying 95% confidence interval. All analyses and data visualization were performed in R version 3.6.1 (21) supplemented with the following packages; MRCIEU/TwoSampleMR (22), rondolab/MR-PRESSO (20), metafor (23), ggplot2 (24).

## Results

### Participant characteristics

Baseline characteristics of the UKB participants are displayed in **Table 1**. Of the 445,482 participants included in this study, 241,337 (54.2%) were women. The mean age□±□SD was 56.8□±□8.0 years, which was similar in men (57.0□±□8.1) and women (56.6□±□7.9). Men were more frequently current smokers (12.2%) than women (8.9%), and more likely to drink alcoholic beverages more than twice a week (53.1% of men, 38.1% of women). Among women, there was a higher prevalence of both a self-reported clinical diagnosis of hypothyroidism (7.7%) and hyperthyroidism (1.2%) compared to the respective 1.6% and 0.3% among men. In line, a higher use of T4 suppletion was reported in women (8.8%) than in men (1.9%), as was the reported use of T3 suppletion and anti-thyroid medication. Mean Hb level□±□SD was 15.0 ± 1.0 g/dL for men and 13.5 ± 1.0 g/dL for women. A total of 18,717 (4.2%) of participants had anemia; 5,907 (2.9%) were men and 12,810 (5.3%) women.

**Table 1.** Participant characteristics at the baseline visit of the UK Biobank

|  | **All** (N = 445,482) | **Men** (N = 204,145) | **Women** (N = 241,337) |
| --- | --- | --- | --- |
| Age in years (Mean ± SD) | 56.8 ± 8.0 | 57.0 ± 8.1 | 56.6 ± 7.9 |
| Smoking currently (n (%)) | 46,412 (10.5%) | 24,903 (12.2%) | 21,509 (8.9%) |
| Alcohol intake >2 times/week (n (%)) | 200,322 (45.0%) | 108,334 (53.1%) | 91,988 (38.1%) |
| hs-CRP (Median (IQR)) | 1.33 (0.66-2.75) | 1.28 (0.66-2.55) | 1.37 (0.65-2.95) |
| GRS TSH (Mean ± SD) | 3.11 ± 0.32 | 3.11 ± 0.32 | 3.11 ± 0.32 |
| GRS fT4 (Mean ± SD) | 2.22 ± 0.21 | 2.22 ± 0.21 | 2.22 ± 0.21 |
| Self-reported clinical diagnosis of thyroid disorder |  |  |  |
| Hypothyroidism (n (%)) | 21,860 (4.9%) | 3,236 (1.6%) | 18,624 (7.7%) |
| Hyperthyroidism (n (%)) | 3,431 (0.8%) | 627 (0.3%) | 2,804 (1.2%) |
| Thyroid hormone suppletion |  |  |  |
| T3 (n (%)) | 123 (0.03%) | 15 (0.01%) | 108 (0.04%) |
| T4 (n (%)) | 25,021 (5.6%) | 3,914 (1.9%) | 21,107 (8.8%) |
| Anti-thyroid medication (n (%)) | 383 (0.09%) | 79 (0.04%) | 304 (0.13%) |
| Hb (Mean ± SD) | 14.19 ± 1.23 | 15.00 ± 1.02 | 13.51 ± 0.95 |
| Anemia (n (%)) | 18,717 (4.2%) | 5,907 (2.9%) | 12,810 (5.3%) |

Abbreviations: BMI; body mass index, GRS; genetic risk score, TSH; thyroid stimulating hormone, fT4; free thyroxine, T4; thyroxine, T3; triiodothyronine, Hb; hemoglobin. Anemia defined as hemoglobin <13 g/dL for men and hemoglobin < 12 g/dL for women. Microcytic anemia mean corpuscular volume (MCV) <80 fL, normocytic anemia MCV 80-100 fL, macrocytic anemia MCV >100 fL.

### Associations between self-reported clinical diagnosis of hypo-and hyperthyroidism with anemia

Associations between self-reported clinical diagnosis of hypo-and hyperthyroidism and anemia are shown in **Table 2**. Self-reported clinical diagnosis of hypothyroidism was associated with higher risk of anemia (OR 1.12, 95% CI 1.05; 1.19, P-value 6.51×10^−4^), independent of C-reactive protein, alcohol intake and smoking. However, the effect estimates of these associations were stronger in men than in women (P-value for interaction 9.48×10^−15^). Though analyses for self-reported clinical diagnosis hyperthyroidism showed a similar direction of effect (odds ratio 1.09), this observation was not supported by evidence from statistical testing (95% CI 0.91; 1.30). Similar to the analyses on self-reported clinical diagnosis of hypothyroidism, the risk estimate was slightly higher in men than in women (P-value for interaction 0.09).

**Table 2.**
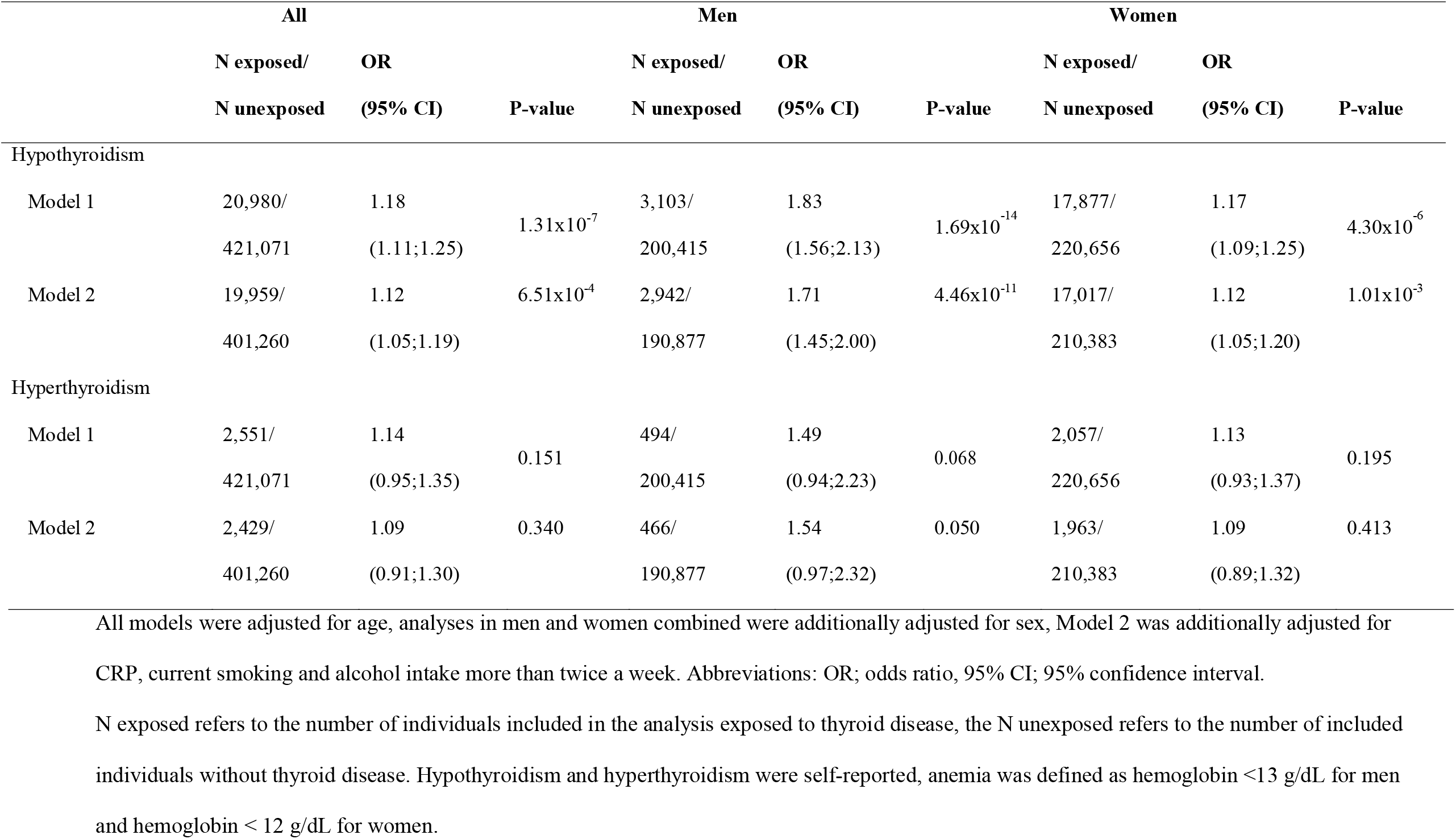
Associations between self-reported clinical diagnosis of hypothyroidism or hyperthyroidism and anemia compared to participants without a self-reported clinical diagnosis of thyroid dysfunction

Results with Hb as a continuous outcome were directionally consistent for hypothyroidism (−0.02 g/dL, 95% CI −0.03; −0.01, P-value 7.09×10^−3^), with a larger effect estimate in men (−0.13 g/dL, 95% CI −0.17; −0.10) than in women (−0.02 g/dL, 95% CI −0.04; −0.01) (**Supplementary table 1**). For hyperthyroidism results were inconsistent with a self-reported clinical diagnosis of hyperthyroidism being associated with higher Hb in all (0.04 g/dL, 95% CI 0.01; 0.08, P-value 0.03) and in women (0.05 g/dL, 95% CI 0.00; 0.08), while with lower Hb in men (−0.03 g/dL, 95% CI −0.13; 0.06).

### Genetically-influenced TSH and fT4 in relation to the risk of anemia

For the MR analyses, TSH and fT3 levels were instrumented in weighted genetic risk scores based on effect estimates of individuals SNPs derived from previous genetic meta-analyses conducted in euthyroid European participants (14). In general, genetically-influenced higher TSH was not associated with risk of anemia. Furthermore, neither individuals with lower genetically-influenced TSH nor those with higher genetically-influenced TSH (expressed as the lower and higher third of the TSH genetic risk score) had an increased risk of having anemia (compared to the middle tertile: OR lowest tertile 0.98, 95% CI 0.95-1.02; highest tertile 1.02, 95% CI 0.98-1.06) (**Table 3**). In addition, genetically-influenced fT4 above or below the reference group was not associated with anemia risk (compared to the middle tertile: OR lowest tertile 1.00, 95% CI 0.96-1.03; highest tertile 0.99, 95% CI 0.95-1.03). Similar results were observed for men and women. Sensitivity analyses on genetically-influenced TSH or fT4 in relation to Hb did not yield any associations either (**Supplementary table 2**). Although some associations between genetically-influenced fT4 and Hb were observed in either men or women only, effect estimates were very small (men with fT4 below average −0.01 g/dL, 95% CI −0.02; 0.00, women with fT4 above average 0.01 g/dL, 95% CI 0.00; 0.02).

**Table 3.** Genetically-influenced thyroid status of TSH and fT4 and anemia in the UK Biobank population^a^

|  | **All** |  | **Men** |  | **Women** |  |
| --- | --- | --- | --- | --- | --- | --- |
|  | **OR (95% CI)** | **P-value** | **OR (95% CI)** | **P-value** | **OR (95% CI)** | **P-value** |
| *Genetically-influenced TSH* |  |  |  |  |  |  |
| Lowest tertile | 0.98 (0.95;1.02) | 0.427 | 1.03 (0.96;1.10) | 0.417 | 0.96 (0.92;1.01) | 0.125 |
| Middle tertile | Reference |  | Reference |  | Reference |  |
| Highest tertile | 1.02 (0.98;1.06) | 0.247 | 1.04 (0.97;1.11) | 0.243 | 1.01 (0.97;1.06) | 0.534 |
| *Genetically-influenced fT4* |  |  |  |  |  |  |
| Lowest tertile | 1.00 (0.96;1.03) | 0.835 | 1.03 (0.96;1.10) | 0.418 | 0.98 (0.94;1.03) | 0.434 |
| Middle tertile | Reference |  | Reference |  | Reference |  |
| Highest tertile | 0.99 (0.95;1.03) | 0.649 | 0.99 (0.92;1.06) | 0.711 | 0.99 (0.95;1.04) | 0.765 |

All analyses were adjusted for age and the first 10 principal components to correct for possible population stratification; analyses in men and women combined were additionally adjusted for sex. Abbreviations: OR; odds ratio, 95% CI; 95% confidence interval.
<sup>a</sup>Anemia defined as hemoglobin <13 g/dL for men and hemoglobin < 12 g/dL for women

Sensitivity analyses using summary-level data-based methods for MR showed similar results, and the MR Egger intercepts and MR PRESSO distortion tests did not indicate presence of severe unbalanced directional pleiotropy except possibly for fT4 and anemia (distortion test p=0.04) (**Supplementary table 3, Supplementary figure 1 to 4**).

### Genetic variation in deiodinases and anemia

Individuals who had genetic variants in *DIO3OS* which were associated with fT4 above or below average had a slightly higher risk of anemia than those in the average group (OR lowest tertile 1.04, 95% CI 1.00-1.08; highest tertile 1.05, 95% CI 1.02-1.10) (**Table 4**). The individual SNPs in *DIO3OS* had similar effect estimates on risk of anemia, though the width of the confidence interval differed (**Supplementary table 4**). No associations were observed with genetic variation in *DIO1* and *DIO2*. In sensitivity analyses, genetic variation in *DIO1, DIO2* and *DIO3OS* was not associated with Hb (**Supplementary table 5**).

**Table 4.**
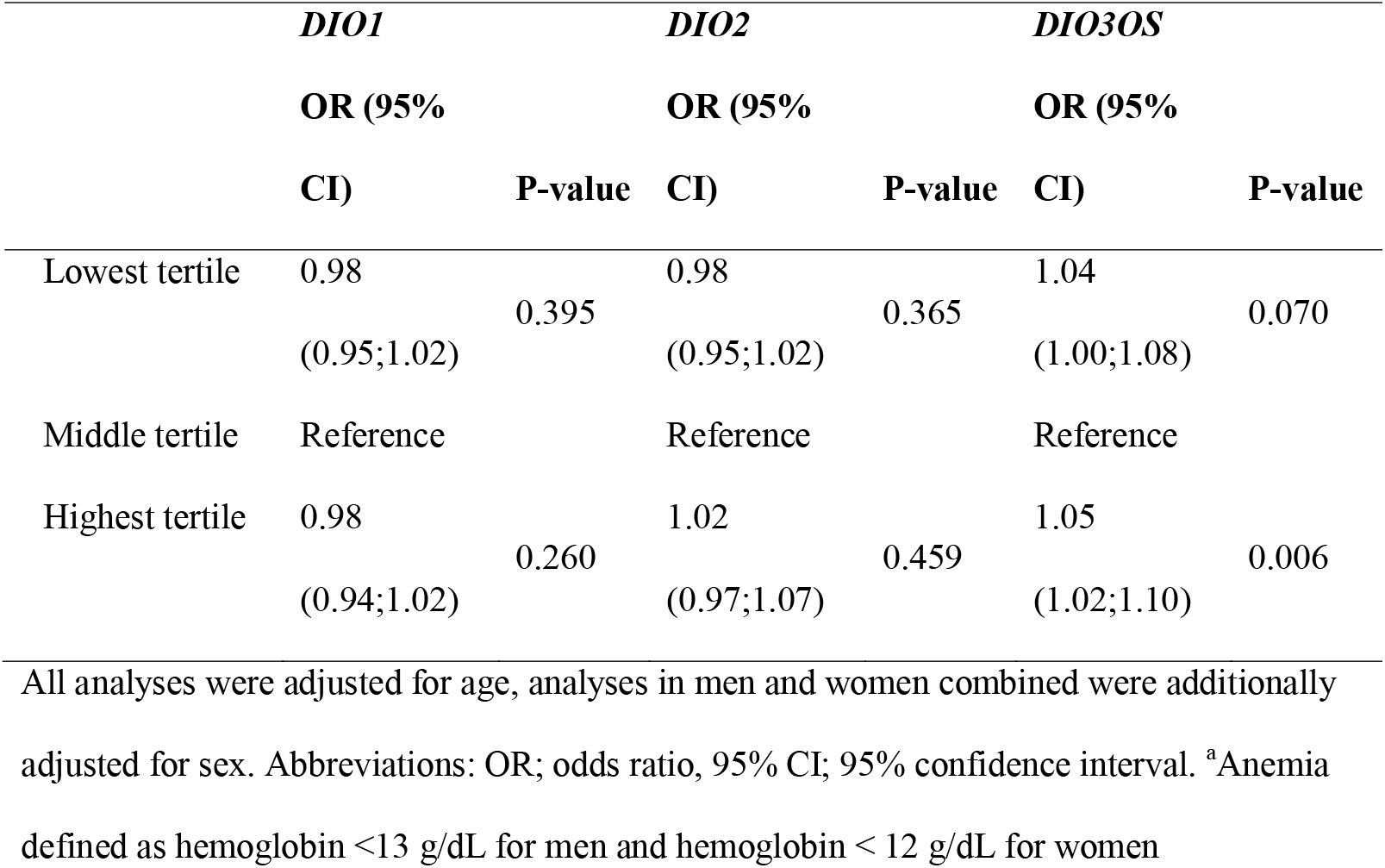
Genetic variation in deiodinase genes and anemia in the UK Biobank population^a^

|  | *DIO1* |  | *DIO2* |  | *DIO3OS* |  |
| --- | --- | --- | --- | --- | --- | --- |
|  | OR (95%<br>CI) | P-value | OR (95%<br>CI) | P-value | OR (95%<br>CI) | P-value |
| Lowest tertile | 0.98<br>(0.95;1.02) | 0.395 | 0.98<br>(0.95;1.02) | 0.365 | 1.04<br>(1.00;1.08) | 0.070 |
| Middle tertile | Reference |  | Reference |  | Reference |  |
| Highest tertile | 0.98<br>(0.94;1.02) | 0.260 | 1.02<br>(0.97;1.07) | 0.459 | 1.05<br>(1.02;1.10) | 0.006 |

All analyses were adjusted for age, analyses in men and women combined were additionally adjusted for sex. Abbreviations: OR; odds ratio, 95% CI; 95% confidence interval. <sup>a</sup>Anemia defined as hemoglobin <13 g/dL for men and hemoglobin < 12 g/dL for women

## Discussion

In this study, multivariable-adjusted and Mendelian Randomization analyses were performed in a large population of individuals of European descent to examine a possible relationship between thyroid function and anemia. We especially found that individuals in our study population with a self-reported clinical diagnosis of hypothyroidism had a higher risk of anemia compared to those without thyroid diseases, especially in men. However, we observed no evidence favoring an association between genetically-influenced variation in circulating TSH and fT4 levels and anemia, suggesting the observed association between the self-reported clinical diagnosis of hypo-or hyperthyroidism with anemia is not causal. Though, in explorative analyses specifically looking at variation in genes encoding deiodinases, a novel significant U-shaped association between variants in the *DIO3OS* gene and risk of anemia was found.

A higher risk of anemia in relation to thyroid diseases has been described previously. In the EPIC-Norfolk cohort, overt hypo-and hyperthyroidism were associated with higher risk of anemia while subclinical thyroid dysfunction was not (11,12). In an individual participant meta-analysis of sixteen cohorts, hypo-and hyperthyroidism were associated with higher prevalence of anemia, and the risk was higher for those with overt thyroid dysfunction than in subclinical disease (10). In the present study, we observed similar results through self-reported data on hypothyroidism, and showed that these associations are independent of (low-grade) systemic inflammation and related lifestyle factors. Adjustment for these factors did not explain the observed difference in association between men and women; It is currently unclear which mechanisms/factors contribute to the higher thyroid-dysfunction-associated anemia risk in men.

Although this is the first study using Mendelian randomization to study genetically-influenced TSH and fT4 levels in relation to anemia, various studies did assess this association using other research designs. Two previous cross-sectional studies also did not observe an association between circulating TSH and Hb levels in euthyroid populations, though both did find an association between low fT4 and lower Hb (25,26). In our analyses, individuals with lower genetically-influenced fT4 level also had lower Hb, but the effect estimates were very small. No association was found between genetically-influenced fT4 and risk of anemia. These differences in findings could be due to residual confounding in the previous studies, or due to imprecision (e.g., lower statistical power, measurement error) of the genetic instruments used in our study. Given that the effect estimates in the Mendelian Randomization analyses were close to zero, it is more likely that residual confounding influences the results from multivariable-adjusted analyses, but additional studies are required to confirm this hypothesis.

To the best of our knowledge, this is the first study to report a significant U-shaped association between genetic variants in the *DIO3OS* gene and anemia risk. Although the exact biological function of *DIO3OS* remains thus far unclear, it is hypothesized that it may be involved in *DIO3* expression levels (27). Temporary induction of D3 induces stem cell proliferation (28), which is also required in erythropoiesis. However, no literature specifically on the role of D3 in erythrocytes or erythropoiesis was found.

Different from the thyroid hormone levels in blood, thyroid hormone deactivation by D3 might affect the risk of anemia. These findings may appear contradictory at first sight, though we hypothesize that they indicate a crucial role for local regulation of thyroid hormone availability in the development of disease. Thyroid signaling can be customized at cellular level through regulation of transporters, deiodinases, amongst others (6). The intracellular exposure to T3 is thereby to some extent independent of centrally regulated TSH and fT4, which is especially important in orchestrating proliferation and differentiation of stem cells (7). Because of this local regulation, processes which are sensitive to thyroid hormone levels might be protected from subtle variations in circulating levels. However, when circulating thyroid hormone levels are extremely high or extremely low these local compensatory mechanisms might no longer suffice, as illustrated previously by Bassett et al. (29). In line with this, when the machinery for regulation does not work optimally (i.e., because of a polymorphism in *DIO2*), local regulation is impaired and cells become more sensitive to changes in circulating levels of thyroid hormones (30). Based on our results, we now hypothesize that functional genetic variants in *DIO3OS* result in a higher risk of anemia driven by a suboptimal intracellular regulation of thyroid hormone inactivation, although this hypothesis requires additional work and functional validation.

The current study has a few noteworthy strengths. First of all, the large sample size of the UK Biobank results in precise effect estimation and allows for stratified analyses even with rare exposures such as hyperthyroidism. Secondly, the combination of different approaches shed light on multiple facets of thyroid function in relation to development of anemia. Together, these new insights add to the etiological understanding of the relationship between thyroid function and anemia. There are also some limitations to the present study. There may be a degree of selection bias, as individuals who responded to the UK Biobank invitation and attended the assessments are healthier than the general population (16). Furthermore, the clinical diagnosis of hypo-and hyperthyroidism were based on self-report; although the interview was conducted by a trained research nurse some misclassification cannot be excluded. Moreover, observed associations of self-reported hypo-or hyperthyroidism with anemia may have been suffered from attenuation due to normalization caused by treatment. Anemia was measured objectively in blood samples taken at the visit, however variation in blood count caused by laboratory drift cannot be ruled out as hematological assays were performed throughout the recruitment period (31). However, this has likely not played a role in the analyses we present in our studies given that this increased variation is most likely caused independently from thyroid function. At last, current analyses were restricted to participants of European ancestry, limiting the generalizability of results to other ancestral groups.

In summary, among individuals of European ancestry participating in UK Biobank, hypothyroidism was associated with a higher risk of anemia independent of inflammation and lifestyle. Genetically determined variation in circulating levels of TSH and fT4 within the reference range is not associated with anemia, though intracellular regulation of thyroid hormones via *DIO3OS* might play a role in development of anemia. Further studies are required to unravel the molecular mechanisms underlying the complex relationship between thyroid hormones and anemia risk.

## Data Availability

Data is open source, and can be accessed after approval of a research proposal to the UK Biobank Review Board and payment of an access fee.

http://www.ukbiobank.ac.uk

## Acknowledgements

This research has been conducted using the UK Biobank Resource under application number 32743. We thank the study participants of the UKB for their participation.

## Funding

This work was supported by the European Commission project THYRAGE (Horizon 2020 research and innovation program, 666869).

## Conflict of interest statement

All authors declare to have no conflict of interest.

## Notes

### Competing Interest Statement

The authors have declared no competing interest.

### Author Declarations

This research has been conducted using the UK Biobank Resource under application number 32743. The UK Biobank has approval from the NHS North West Multi-Centre Research Ethics Committee (ref 11/NW/0382). All participants from the UK Biobank cohort provided written informed consent, and the study was approved by the medical ethics committee.

