## Supplementary Materials for "Thyroid function and risk of anemia: a multivariable-adjusted and Mendelian Randomization analysis in the UK Biobank"

### **Supplementary table 1.** Associations between self-reported clinical diagnosis of hypothyroidism or hyperthyroidism and hemoglobin compared to participants without a self-reported clinical diagnosis of thyroid dysfunction

|  | **All** | | | **Men** | | | **Women** | | |
| --- | --- | --- | --- | --- | --- | --- | --- | --- | --- |
|  | **N exposed/**  **N unexposed** | **Beta**  **(95% CI)** | **P-value** | **N exposed/**  **N unexposed** | **Beta**  **(95% CI)** | **P-value** | **N exposed/**  **N unexposed** | **Beta**  **(95% CI)** | **P-value** |
| Hypothyroidism |  |  |  |  |  |  |  |  |  |
| Model 1 | 20,980 /  421,071 | -0.02  (-0.03;-0.01) | 3.18x10^-3^ | 3,103 / 200,415 | -0.14  (-0.17;-0.10) | 1.81x10^-13^ | 17,877 / 220,656 | -0.02  (-0.04;-0.01) | 2.16x10^-3^ |
| Model 2 | 19,959/  401,260 | -0.02  (-0.03;-0.01) | 7.09x10^-3^ | 2,942/  190,877 | -0.13  (-0.17;-0.10) | 2.20x10^-12^ | 17,017/  210,383 | -0.02  (-0.04;-0.01) | 4.02x10^-3^ |
| Hyperthyroidism |  |  |  |  |  |  |  |  |  |
| Model 1 | 2,551 /  421,071 | 0.05  (0.01;0.09) | 0.013 | 494 /  200,415 | -0.02  (-0.11;0.06) | 0.587 | 2,057 /  220,656 | 0.05  (0.01;0.09) | 0.018 |
| Model 2 | 2,429/  401,260 | 0.04  (0.01;0.08) | 0.025 | 466/  190,877 | -0.03  (-0.13;0.06) | 0.474 | 1,963/  210,383 | 0.04  (0.00;0.08) | 0.048 |

All models were adjusted for age, analyses in men and women combined were additionally adjusted for sex,

Model 2 was additionally adjusted for CRP, current smoking and alcohol intake more than twice a week. Abbreviations: 95% CI; 95% confidence interval.

N exposed refers to the number of individuals included in the analysis exposed to thyroid disease, the N unexposed refers to the number of included individuals without thyroid disease. Hypothyroidism and hyperthyroidism were self-reported, beta represents difference in Hb in g/dL.

### **Supplementary table 2**. Genetically determined thyroid status of TSH and fT4 and hemoglobin levels in the UK Biobank population

|  | **All** | | **Men** | | **Women** | |
| --- | --- | --- | --- | --- | --- | --- |
| **Genetic Risk Score** | **Beta (95% CI)** | **P-value** | **Beta (95% CI)** | **P-value** | **Beta (95% CI)** | **P-value** |
| *TSH* |  |  |  |  |  |  |
| Lowest tertile | 0.000 (-0.007; 0.008) | 0.991 | -0.003 (-0.014; 0.008) | 0.573 | 0.003 (-0.007; 0.013) | 0.559 |
| Middle tertile | Reference |  | Reference |  | Reference |  |
| Highest tertile | -0.004 (-0.012; 0.003) | 0.290 | -0.002 (-0.014; 0.009) | 0.682 | -0.006 (-0.015; 0.004) | 0.264 |
| *fT4* |  |  |  |  |  |  |
| Lowest tertile | -0.007 (-0.014; 0.001) | 0.07 | -0.011 (-0.023; 0.000) | 0.049 | -0.003 (-0.013; 0.006) | 0.500 |
| Middle tertile | Reference |  | Reference |  | Reference |  |
| Highest tertile | 0.007 (0.001; 0.014) | 0.08 | 0.003 (-0.009; 0.014) | 0.640 | 0.010 (0.001; 0.020) | 0.038 |

All analyses were adjusted for age, analyses in men and women combined were additionally adjusted for sex. Abbreviations: 95% CI; 95% confidence interval.

**Supplementary table 3.** Sensitivity analyses for genetically determined thyroid status and anemia and hemoglobin levels

|  | **Anemia** | | **Haemoglobin** | |
| --- | --- | --- | --- | --- |
|  | **logOdds (SE)** | **P-value** | **Beta (SE)** | **P-value** |
| ***Genetically-influenced TSH*** |  |  |  |  |
| IVW | 0.017 (0.042) | 0.68 | -0.006 (0.019) | 0.76 |
| WME | -0.049 (0.049) | 0.33 | 0.011 (0.010) | 0.28 |
| MR Egger |  |  |  |  |
| Estimate | 0.086 (0.103) | 0.41 | -0.047 (0.046) | 0.32 |
| Intercept | -0.005 (0.007) | 0.47 | 0.003 (0.003) | 0.34 |
| MR-PRESSO |  |  |  |  |
| Global test | . | 0.008 | . | <0.001 |
| Distortion test | . | 0.31 | . | 0.27 |
| ***Genetically-influenced fT4*** |  |  |  |  |
| IVW | 0.009 (0.076) | 0.91 | 0.018 (0.042) | 0.66 |
| WME | -0.009 (0.065) | 0.89 | 0.020 (0.014) | 0.14 |
| MR Egger |  |  |  |  |
| Estimate | 0.175 (0.180) | 0.34 | -0.055 (0.099) | 0.59 |
| Intercept | -0.013 (0.013) | 0.32 | 0.006 (0.007) | 0.43 |
| MR-PRESSO |  |  |  |  |
| Global test | . | <0.001 | . | <0.001 |
| Distortion test | . | 0.04 | . | 0.22 |

Abbreviations: IVW; Inverse-variance weighted, WME; Weighted Median Estimator, MR-PRESSO; Mendelian randomization pleiotropy residual sum and outlier, TSH; thyroid stimulating hormone, fT4; free thyroxine.

**Supplementary table 4.** Variation in deiodinase genes and heamoglobin

|  | ***DIO1*** | | ***DIO2*** | | | ***DIO3OS*** | |
| --- | --- | --- | --- | --- | --- | --- | --- |
|  | **Beta (95% CI)** | **P-value** | | **Beta (95% CI)** | **P-value** | **Beta (95% CI)** | **P-value** |
| Lowest tertile | -0.001  (-0.010; 0.006) | 0.785 | | -0.004  (-0.003; 0.010) | 0.252 | -0.003  (-0.010; 0.005) | 0.504 |
| Middle tertile | Reference |  | | Reference |  | Reference |  |
| Highest tertile | 0.003  (-0.005; 0.011) | 0.474 | | -0.009  (-0.002; 0.019) | 0.098 | -0.001  (-0.009; 0.006) | 0.741 |

All analyses were adjusted for age, analyses in men and women combined were additionally adjusted for sex. Abbreviations: OR; odds ratio, 95% CI; 95% confidence interval.

**Supplementary Table 5.** Genetically determined thyroid status of separate DIO3OS SNPs and anemia

|  | **rs11626434** | | **rs12323871** | |
| --- | --- | --- | --- | --- |
|  | **OR (95% CI)** | **P-value** | **OR (95% CI)** | **P-value** |
| Lowest tertile | 1.03 (1.00-1.06) | 0.075 | 1.03 (0.97-1.11) | 0.345 |
| Middle tertile | Reference |  | Reference |  |
| Highest tertile | 1.04 (1.00-1.09) | 0.064 | 1.03 (0.96-1.10) | 0.403 |

All analyses were adjusted for age and sex. Abbreviations: OR; Odds Ratio, 95% CI; 95% confidence interval.

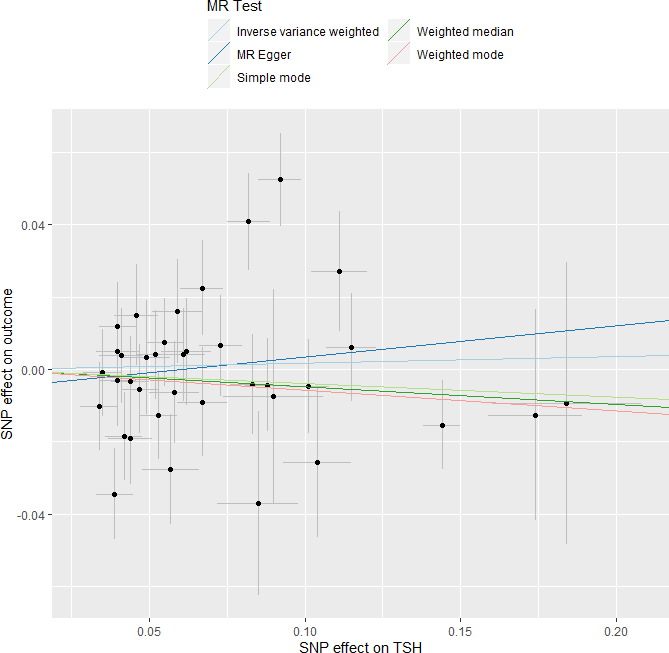

**Supplementary figure 1.** SNP effect on TSH and anemia

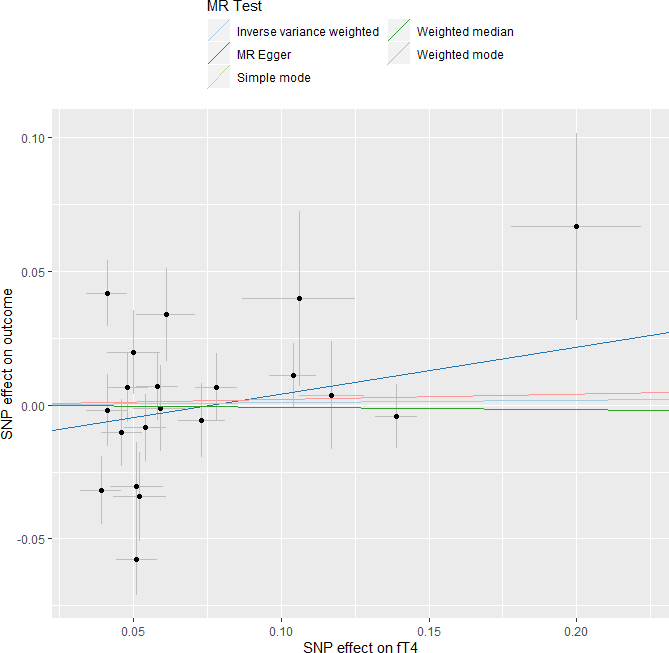

**Supplementary figure 2.** SNP effect on fT4 and anemia

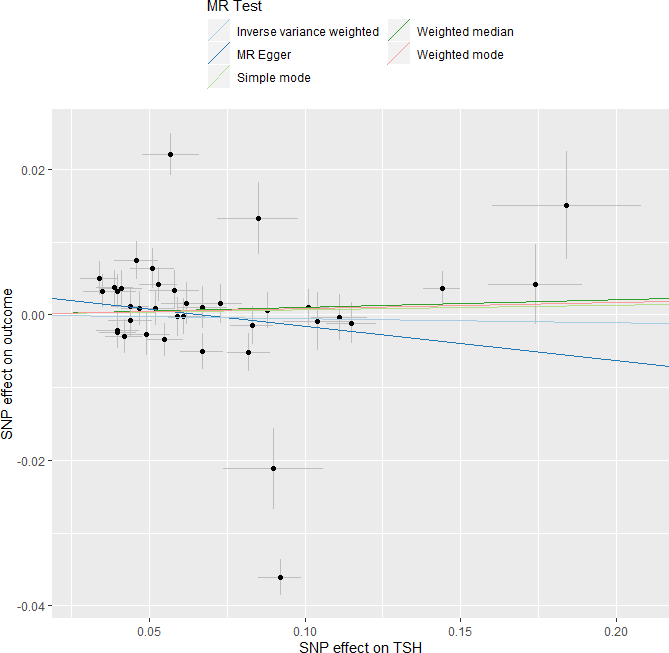

### **Supplementary figure 3.** SNP effect on TSH and Hb

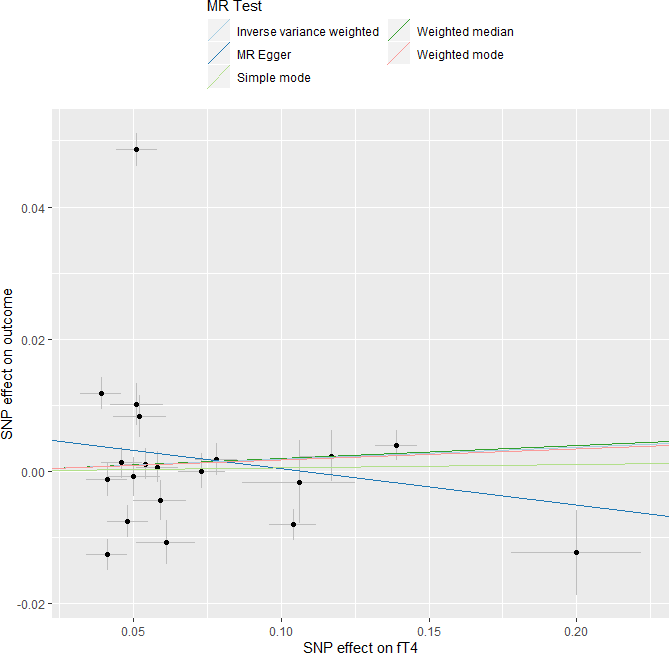

### **Supplementary figure 4.** SNP effect on fT4 and Hb
